# Human fingertip regeneration follows clinical phases with distinct proteomic signatures – a Comprehensive MS/MS analysis

**DOI:** 10.1101/2025.06.06.25329128

**Authors:** Jurek Schultz, Purva A. Patel, Rita Aires, Leah Wissing, Patrick Glatte, Michael Seifert, Marc Gentzel, Guido Fitze, Adele M. Doyle, Tatiana Sandoval-Guzmán

## Abstract

Distal injuries in human fingertips can regenerate almost fully, yet the process of human fingertip regeneration has hardly been characterized on a cellular and molecular level. A silicone finger cap, comprising a puncturable reservoir, was used to treat 22 human fingertip amputations. In all patients, subcutaneous tissue, nailbed and skin regenerated with excellent outcomes. Through the clinical assessment of the wounds, the regenerative process was divided into four distinct phases. Proteomic data from wound fluid samples collected at regular intervals, confirmed robust and unbiasedly distinct proteomic signatures, characteristic processes, and active regulatory networks in each phase. Moreover, this human dataset provides important insights, showing clear divergences from findings in regenerative animal models. The longitudinal and comprehensive analysis presented here unveils the complex orchestration of four clinically and proteomically-distinct phases of human fingertip regeneration. Further analyses of this proteomic data will allow for the identification of candidates orchestrating human fingertip regeneration and serving as a framework for comparative and regenerative medicine studies.

## Main

The fingertips are of utmost importance for our tactile perception of the world^1^. Without their extremely fine sensitivity and, simultaneously, their robust nature, the functionality of the hand is severely impaired^2^. At the same time, the cosmetic appearance of the hands is equally important for many patients^3^. Because of their exposed position and critical role in most human activities, fingertip injuries are extremely common. Their annual incidence in the USA has been reported to be 7.5/100000^1^ and account for more than 4.8 million ER presentations per year^4^. Surgical approaches for fingertip amputation injuries unsuitable for primary closure include stump plasties, local or distant flaps, microsurgical replantation, composite grafts, or skin transplants^5^. However, non-operative management has long been demonstrated to have superior outcomes compared to surgical techniques^6–8^. This superiority is mainly due to the *de novo* formation of subcutaneous tissues and epithelium, rather than tissue contraction^9^.

The extraordinary capacity of the human distal fingertip to regenerate is evident in the restoration of fingerprints and scarless regeneration^10,11^. This regeneration is facilitated under semi-occlusive dressings that keep hydration, temperature, pH, oxygen saturation, and other environmental values in favorable ranges while retaining potentially pro-regenerative factors within the wound fluid^12,13^. In contrast, when the wound dries out, a failure to regenerate and severe scarring can be observed^14^. The extraordinary regenerative outcome of the human fingertip, including recovery of the original shape and appearance, places it as possibly the only example of epimorphic regeneration in humans.

Examples of limb regeneration in the animal kingdom are found in broadly-studied models like urodele amphibians (salamanders), which show a fascinating ability to regenerate during their entire lifespan^15–17^. Even after a proximal amputation, they can restore the complete missing limb^18^. Limb regeneration is achieved through epimorphosis, which is defined as regeneration through extensive cell proliferation and morphogenesis, and through the formation of a temporal structure supporting the massive initial growth, the blastema^19^. A well-defined characteristic of limb regeneration in salamanders is the rapid wound closure and re-epithelialization within the first hours post-injury, a stark difference from mammalian wound healing. Regeneration in mammalian animal models is restricted to the distal part of the fingertips (P3-segment)^20,21^. Particularly, regeneration in the mouse terminal phalanx is a well-established model^22,23^, and can be divided into specific phases. After an initial phase of coagulation, inflammation, and histolysis, a wound epidermis forms, closing the amputation injury (wound healing). Similar to urodeles, the blastema forms under the wound epidermis (blastemal formation and growth) to regrow the missing fingertip, including the bone (differentiation and morphogenesis)^22,24,25^.

However, insights into human fingertip regeneration (HFR) are limited compared to animal models. A few examples include clinical reports and studies on fetal tissues^26^. Given that both human wound healing and epimorphic limb regeneration in animal models have been described to follow sequential but overlapping phases^23,27^, we hypothesized a similar sequence in HFR. In addition to the insufficient characterization of the clinical phases of HFR, there is also little evidence on the mechanisms that drive HFR^26,28^. Therefore, for the first time, we studied HFR in a randomized controlled clinical trial, including the review of patients records, clinical photographs, x-rays, and a comprehensive analysis of the proteome detected in the wound fluid collected during the course of HFR. We defined the phases of HFR and characterized their timing depending on the severity of the initial injury. In line with clinical reports^12,28,29^, we noticed characteristic macroscopic features during fingertip regeneration in many patients^30^. We show with unbiased approaches that distinct clinical and proteomic phases are consistent across the cohort of patients, with no apparent correlation to the heterogeneity of injury severity, injury nature, sex, age and timing of sample collection. These robust phases, defined by the macroscopic features and proteomes, are expected to be relevant for future studies on human regeneration.

## Results

### HFR follows a typical sequence of events grouped into 4 phases

Through a comprehensive analysis of all available patient data, we defined the following clinical phases of human fingertip regeneration: 1) Coagulation: Bleeding ceases spontaneously, and within one day, a blood clot covers the amputation site. 2) Hypergranulation: During the following days, granulation tissue forms and covers the wound. This granulation tissue slowly grows into and eventually even exceeds the former limits of the original fingertip. 3) Proliferation: Then, a keratinized epithelium begins to cover the hypergranulation tissue starting at the proximal wound edge. 4) Epithelialization: At the end of this process, the regenerated tissues are nearly covered by the new epithelium. In some instances, distal parts of the hypergranulation tissue are extruded from the new epithelium. At the most distal part of the regenerated fingertip, some patients may present with a small area not covered by skin. On this case, the extruded hypergranulation tissue eventually dries out and sheds in the form of a scab. The remaining skin gaps close with minimal scarring or normal-looking skin. Representative images of these phases are shown in Fig. 1a. An assessment of the wounds was performed to classify the injuries according to the Allen score described in detail in the methods section. A summary of the age and Allen score is provided in Fig. 1b. To uncover if there was a difference in the regeneration time dependent on the initial severity of the amputation (scored as Allen, II, III or IV), we analyzed the day the treatment ended (regeneration was completed). On average, patients with Allen IV needed 33.6 more days to regenerate than patients with Allen type II injuries and 30.14 more days than patients with Allen III (Fig. 1c). Once the regenerative phases were clinically defined for all patients, accounting for all the visits, the wound fluid samples were assigned to their matching phase (Fig. 1d).

**Fig. 1.**
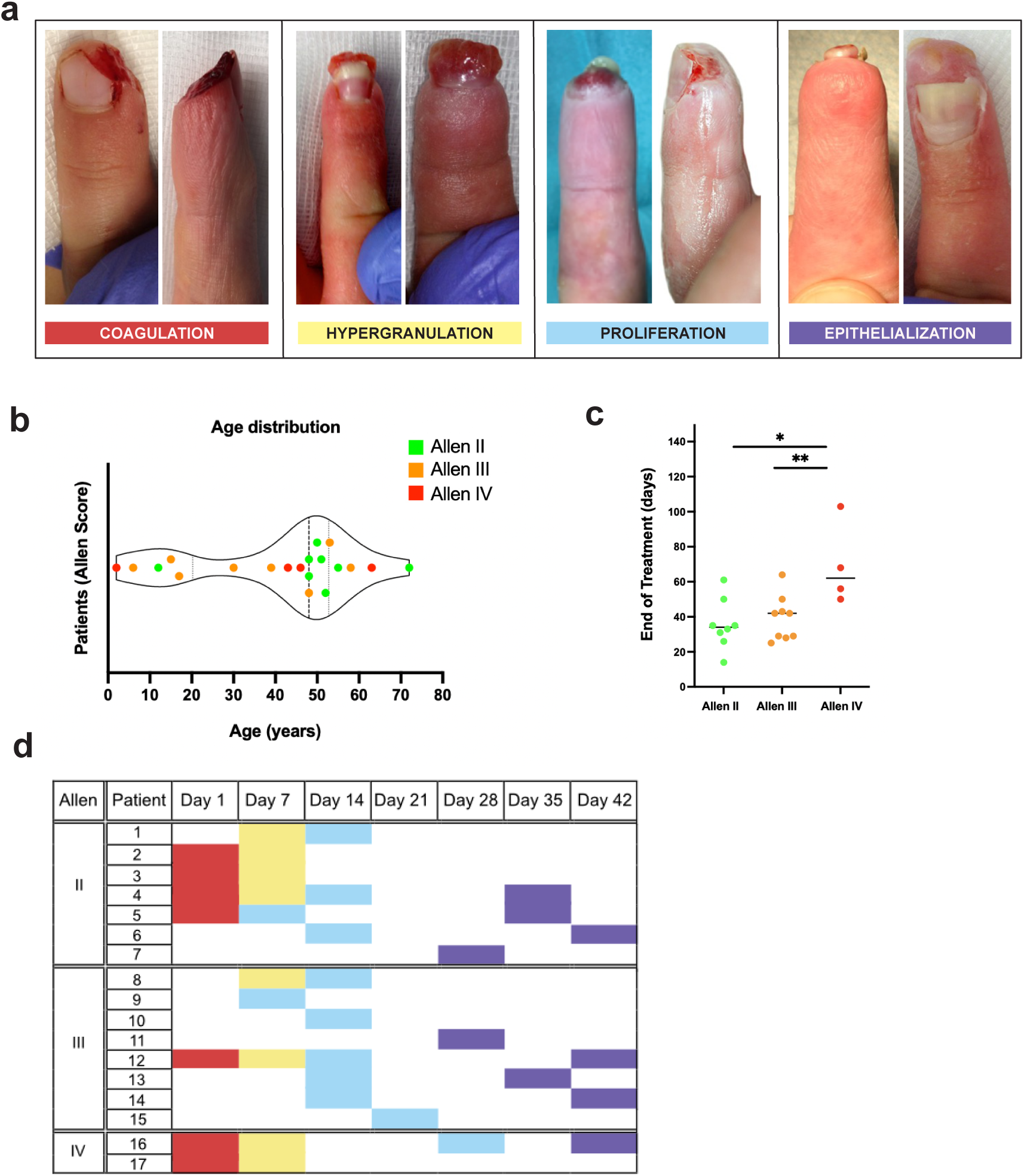
Summary of the cohort and regenerative phases during HFR. **a.** Two representative photographs of each of the four different phases of human fingertip regeneration (HFR). Images were taken during the scheduled trial visits. **b.** Overview of the age distribution and Allen score of the patients in this study. **c**. Mean duration of the treatment by type of injury according to the Allen score. Each dot represents one patient. One-way ANOVA and Tukey’s multiple comparison test. \**p*≤0.05, ** *p*≤0.01. Mean is shown as a line. **d.** Sample distribution of patients included in the MS/MS, classified according to the Allen Score (see Materials and Methods) for fingertip injuries.

### Wound proteomics of the regenerative phases of HFR

Wound fluids were very heterogeneous samples, from liquid to more viscous consistency or with visibly more lipids. However, we analyzed the proteomic composition of 36 samples by MS/MS data from: 7 samples of Coagulation, 8 of Hypergranulation, 12 of Proliferation, and 9 of Epithelialization. We analyzed the total spectrum counts (TSC) for the proteins in each sample to create a basic common protein count data matrix. The protein abundance was analyzed and 974 reviewed proteins were identified in at least one of the 36 samples (Supplementary Data 1). From those, 690 proteins were identified in the Coagulation phase, 788 proteins in the Hypergranulation phase, 797 in the Proliferation phase and 502 in the Epithelialization phase.

To understand how proteins define each regenerative phase, a clustered heatmap of all proteins in all samples was created and represented here by phase in chronological order (Fig. 2a). Total spectrum counts for the proteins in each sample were used to determine if the samples assigned to a clinical phase were distinctly different from each other. 2-way ANOVA test with Tukey’s multiple comparison test revealed that all group comparisons are significantly different from each other (p <0.0001), except for the comparison Coagulation *vs* Hypergranulation (Supplementary Data 2). To further understand the sample variations of the healing phases, we performed a principal component analysis (PCA) on the 36 samples; the 3 highest-weighted components are represented in a 3D scatter plot (Fig. 2b): PC1 68.8%, PC2 5.8%, and PC3 5.8%. In this analysis, the comparison of PC1 versus PC2 captures the separation of phases 1 and 2 versus 4 (Coagulation and Hypergranulation versus Epithelialization) via PC1 and Coagulation vs Epithelialization via PC2, while the plot PC1 versus PC3 (Fig. 2c) captures the variation of phases 1 and 3, Coagulation versus Proliferation in PC3. While the two earliest phases show robust separation versus the latest phase (PC1), the intermediate Proliferation phase shows comparatively slight clustering (PC3) suggesting a similarity to other phases, or alternatively, it reflects the diversity of the growth originated from diverse injuries. PC1 is characterized by the contribution of albumin and hemoglobin as proteins exhibiting the greatest variation during healing. In contrast, PC2 is heavily influenced mainly by KRT10 and Albumin, while PC3 relies on both KRT10 and Hemoglobin (Supplementary Fig. 1).

**Fig. 2.**
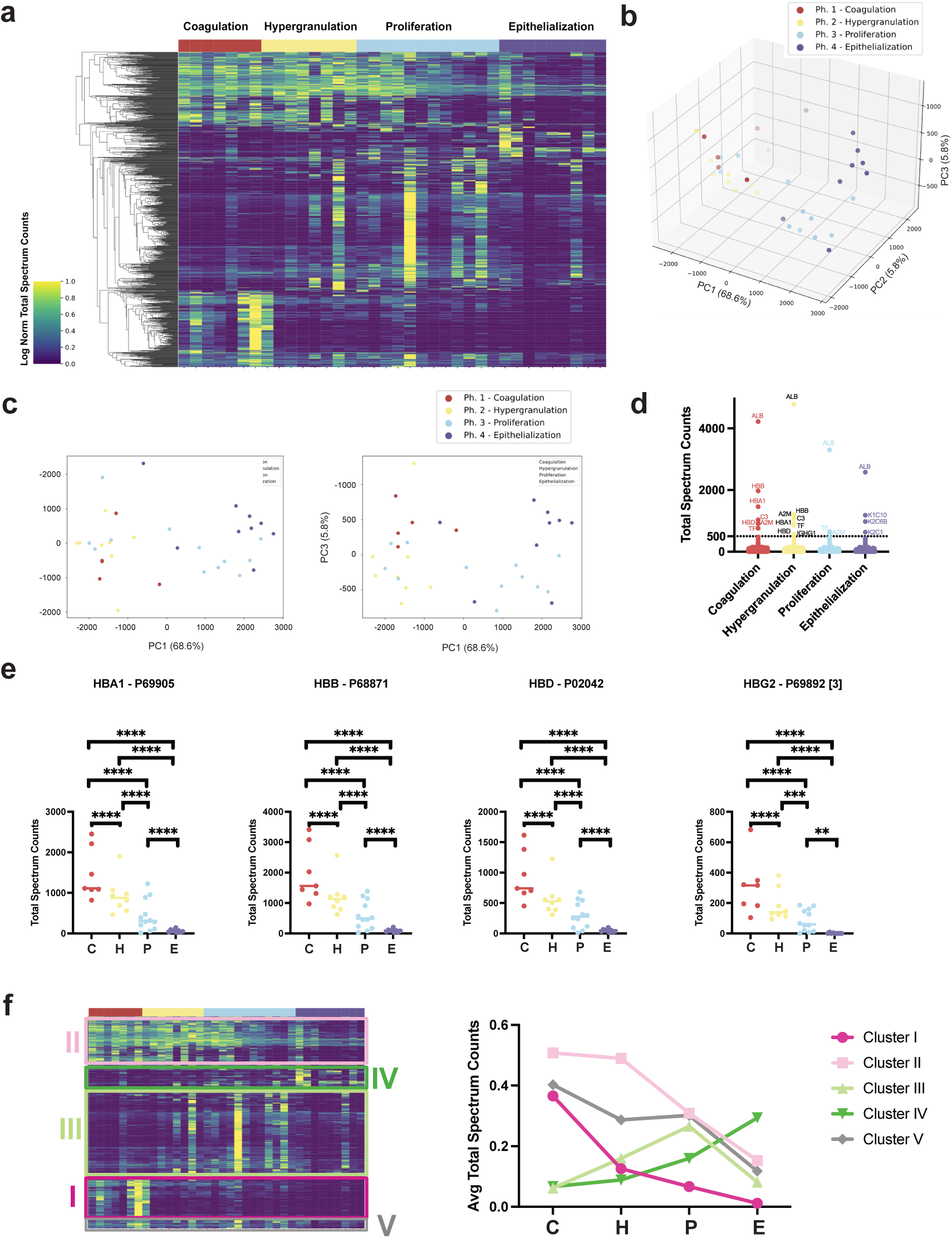
MS/MS proteomic data distinguishes wound fluid samples into 4 phases during HFR. **a**. Protein expression was measured in wound fluids (n=36 samples) using mass spectrometry. Hierarchical clustering of wound samples with each other, aggregates samples into four groups, as shown by matching color blocks for HFR phase assignments made for each sample from clinical data. Linkage method ward. 980 proteins of 2481 total detected were significantly different, clustering in 4 groups of similar expression **b**. Principal component analysis shows the four regeneration phases separate using the first three components. Each dot represents one proteomic measurement; colors mark the regeneration phase. **c**. 2D PCA showing PCA1 vs PCA2 and PCA1 vs PCA3. **d**. Most abundant proteins (Total Spectrum Counts) by phase. **e**. Hemoglobin, a primary contributor to variation in PC2, significantly decreases during HFR for all 8 forms detected by MS/MS analysis. One way ANOVA; significance denoted by ns (*p*>0.05), *(*p*≤0.05), **(*p*≤0.01), ***(*p*≤0.001), or ****(*p*≤0.0001). **f**. Clustered heatmap shown in Fig. 1a, with highlights of subclusters based on expression pattern (left), represented as different colors, and the average total spectrum counts graphed for each cluster (right). The x-axis represents the four phases, the y-axis the average spectrum counts. C= Coagulation, H= Hypergranulation, P= Proliferation, E= Epithelialization.

The most abundant proteins in each regenerative phase are plotted in Fig. 2d, showing albumin to be the most abundant protein across all phases, followed by hemoglobin subunits (HBB, HBA1, HBD). Complement protein C3 is prevalent in the earlier phases, contrasting with Keratins dominating the later Epithelialization phase. Additionally, alpha 2-macroglobulin (A2M), a protease inhibitor, is highly detected in the first 3 phases. Because hemoglobin subunits are highly abundant, we evaluated whether significant differences between phases could be detected. We found that hemoglobin subunits were expressed at high levels in recent wounds while decreasing as regeneration progresses, with statistical significance across the regenerative phases (Fig. 2e). Taken together, these data indicate that wound fluid contains protein-based reporters of the tissue wound response and regeneration, which further cluster based on regeneration phase. To investigate the proteins that contribute to differentiating the regenerative phases, we analyzed distinct clusters of proteins identified by their specific abundances in each phase. This resulted in five identifiable clusters (Fig. 2f, Supplementary Fig. 2). Cluster I comprises a group of proteins that are abundant in Coagulation, but sharply decrease in the subsequent phases. Cluster II proteins also decrease after Coagulation, but remain moderately abundant in subsequent phases. Cluster III peaks in the Proliferation phase. Cluster IV shows a slow increase and a peak at the Epithelialization phase. Lastly, Cluster V is defined by proteins present in the four phases with a moderate decrease. Components of the different clusters can be found in Supplementary Data 3.

### Functional groups of differentially expressed proteins (DEPs)

From this universe of proteins, we analyzed those differentially expressed when comparing their expression to each of the other phases. Differentially detected proteins (differentially expressed proteins, DEPs) were identified using one-factor ANOVA (regeneration phase, determined using clinical assessment) and post-hoc Tukey’s test. Of 974 detected proteins, 60 showed significantly differential expression between two or more regeneration phases (Supplementary Data 4). The normalized average count of these 60 proteins was plotted as a clustered heatmap to highlight their association by abundance across phases (Fig. 3a). The abundance pattern exposes proteins enriched by phase and those that mark transitional expression.

**Fig. 3.**
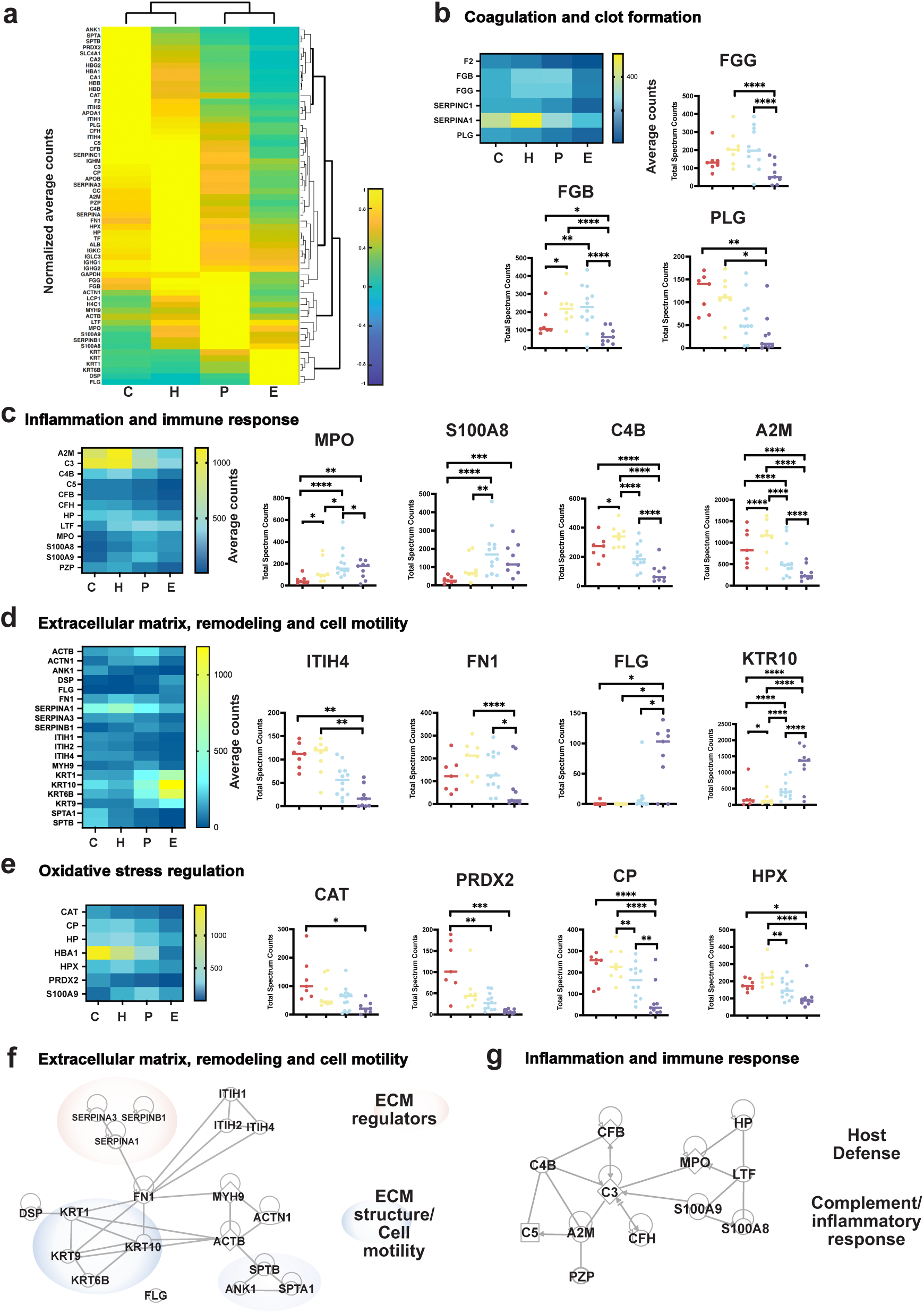
Individual differentially expressed proteins (DEPs) group by phase and by healing function across phases. **a**. Clustered heatmap of the 60 DEPs normalized average spectral counts, columns are arranged by regenerative phase and rows show the associations between the proteins and their abundance by phase. **b**. Heatmap of DEPs associated with coagulation and clot formation, and 3 representative proteins graphed by individual samples per phase. Fibrinogen gamma chain (FGG), Fibrinogen beta chain (FGB), and plasminogen (PLG). **c**. Heatmap of DEPs associated with inflammation and immune response, and 4 representative proteins graphed by individual samples per phase. Myeloperoxidase (MPO), S100 calcium binding protein A8 (S100A8), Complement C4B, (C4B), Alpha 2 macroglobulin (A2M). **d.** Heatmap of DEPs associated with extracellular matrix, remodeling, and cell motility, and 4 representative proteins graphed by individual samples per phase. Inter-alpha-trypsin inhibitor heavy chain 4 (ITIH4), Fibronectin (FN1), Filaggrin (FLG), Keratin 10 (KRT10). **e**. Heatmap of DEPs associated with oxidative stress regulation and 4 representative proteins graphed by individual samples per phase. Catalase (CAT), Peroxiredoxin 2(PRDX2), Ceruloplasmin (CP), and Hemopexin (PHX). **f**. IPA analysis of DEPs in the extracellular matrix, remodeling, and cell motility cluster. Linked genes with direct interactions sub-grouped in ECM regulators (pink clouds) and ECM structural and cell motility components (blue clouds). Fibronectin acts as the connecting protein with direct interactions to all subgroups. **g**. IPA analysis of DEPs in the Inflammation and Immune Response cluster. Linked genes with direct interactions sub-grouped in Host defense (gray cloud) and Complement/inflammatory response (yellow cloud). C3 acts as the connecting protein with direct interactions to all subgroups. For all panels in Fig. 3: The median is represented by a line and each dot is an individual sample. C= Coagulation, H= Hypergranulation, P= Proliferation, E= Epithelialization. \**p*≤0.05, \*\**p*≤0.01, \*\*\**p*≤0.001, \*\*\*\**p*≤0.0001.

Functional grouping of the DEPs resulted in 8 groups. The most populated groups were plotted as heatmaps next to representative proteins of this group, plotted by phase and values of individual samples (Fig. 3b-e). The proteins involved in coagulation and clot formation (F2, FGB, FGG, PLG, SERPINC1, SERPINA1) are prevalent in the first three phases (Fig. 3b). Proteins related to inflammation and immune response (C3, C4B, C5, CFB, S100A8/9, MPO, HP, SERPINA1, LTF, A2M, PZP) subgroup in proteins abundant in the first two phases, while a second subgroup is more abundant in the two middle phases (Fig. 3c). Extracellular matrix remodeling and cell motility proteins (ACTB, ACTN1, ANK1, FN1, ITIH1, ITIH2, ITIH4, DSP, FLG, KRT10, KRT9, KRTC6B, MYH9, SERPINB1, SERPINA3, SERPINA1, SPTA, SPTB) are abundant in the first three phases with a clear switch to keratins in the Epithelialization phase (Fig. 3d). Oxidative stress regulation proteins (HBA1, CAT, PRDX2, CP, HP, HPX) are present mainly in the first three phases (Fig. 3e). In addition, transport proteins form a functional group (TF, APOA1, LTF, ALBUMIN, APOB, CP, GC); metabolic proteins are present predominantly in the first three phases (CA1, CA2, GAPDH, GC, SLC4A1); hemoglobin (HBA1, HBB, HBD, HBG2) in a decreasing trend (Fig. 2e) and immunoglobulins (IGHG1, IGHG2, IGHM, IGKC, IGLC3) in all four phases.

We then used Ingenuity Pathway Analysis (IPA) of the 60 DEPs to identify direct interactions of proteins within sets and protein regulators (Fig. 3f, g). In the group of extracellular matrix, remodeling and cell motility proteins, four groups containing ECM regulators and structural proteins connect through Fibronectin (FN1). In the inflammation and immune response, two subgroups containing proteins related to host defense and Complement/Inflammatory response connect by the protein complement C3. From the 60 DEPs, 31 identify as extracellularly located, 18 as cytoplasmic, 6 in the plasma membrane, 2 in the nucleus, and 3 as other. When all 60 DEPs are included in the IPA network analysis, two main networks appear, one network with a prominent presence of blood proteins and a second network prominent with components of ECM and its regulators (Supplementary Fig. 3). The remaining 7 networks are available in Supplementary Data 5.

### Regenerative phase comparison

To unbiasedly define the differences between the phases, we performed a sequential phase comparison using IPA with all proteins included in all the samples. The Canonical Pathways with a high activation z-score when compared to subsequential phases are predominantly neutrophile degranulation, followed by signaling pathways related to the immune response and cholesterol metabolism (Fig. 4a and Supplementary Data 6). A heatmap of activation diseases and function phase comparison revealed that the top diseases and biofunctions with inhibition scores are associated with cell death (Fig. 4b and Supplementary Data 7). Higher activation scores are dominated by migration of cells, migration of blood cells and metabolism, adhesion of cells and cell survival, followed by terms associated with immune and inflammatory response formation of angiogenesis and vasculogenesis. Next, we plotted the z-scores of the significantly differentiating canonical signaling terms to their respective *p*-values to understand their activation with respect to their significant difference and trajectory through phases. These terms could be grouped in signaling pathways (Fig. 4c), immune response (Fig. 4d), as well as coagulation and clotting (Fig. 4e) (Supplementary Data 8). In the transition from Hypergranulation to Proliferation, the terms with higher significance and activation are complement cascade, acute phase response signaling, neutrophil degradation, LXR/RXR activation and TRIM21 intracellular antibody signaling pathway. For the transition between Proliferation and Epithelialization, some terms overlap with the previous comparisons, such as complement cascade, neutrophil degranulation, response to elevated cytosolic Ca^2+^, acute phase signaling and TRIM21 intracellular antibody signaling pathway (Fig. 4d).

**Fig.4.**
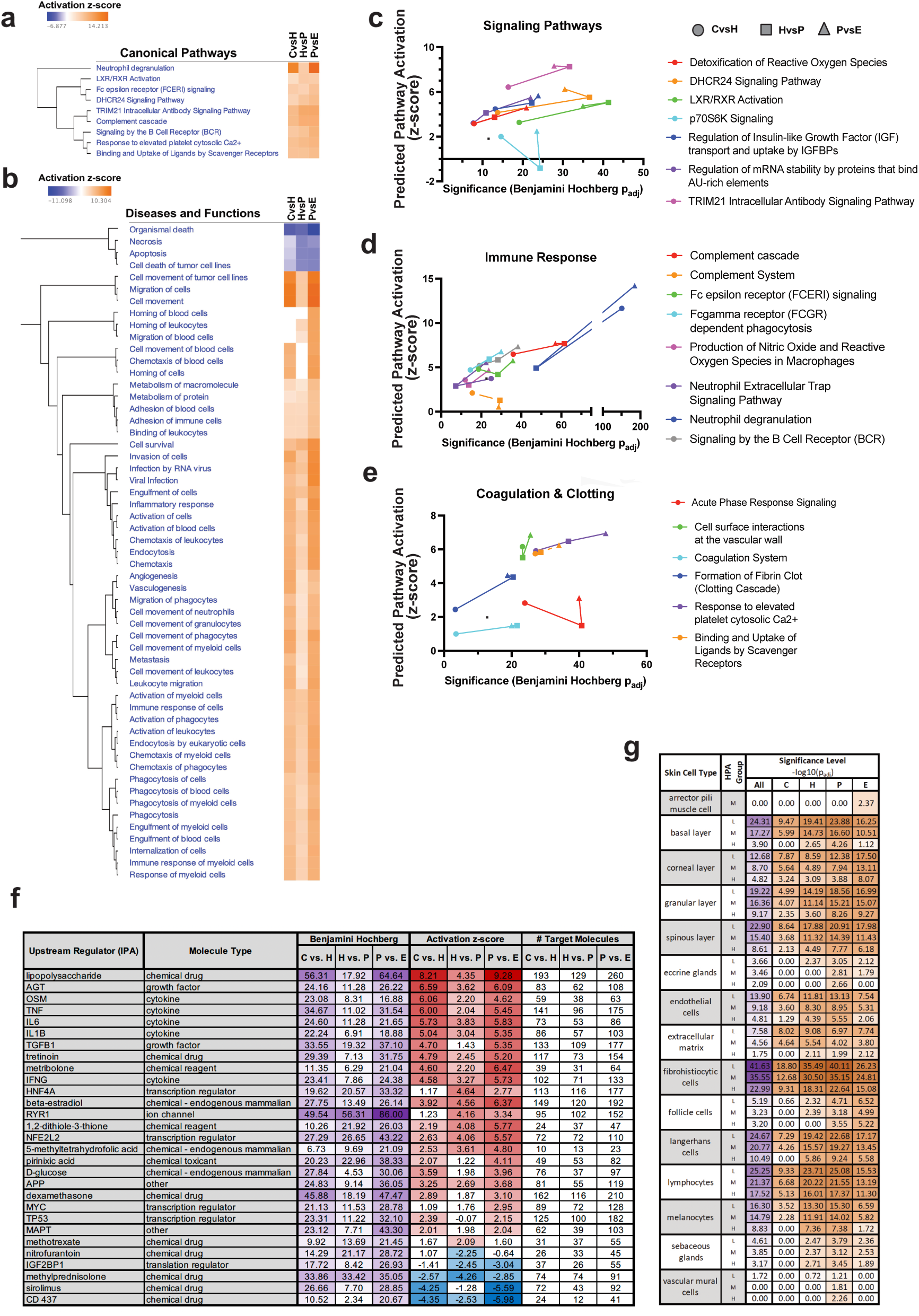
HFR proteome analysis. **a**. Canonical pathways phase comparison analysis. Each column represents a pairwise comparison: Coagulation vs Hypergranulation (CvsH), Hypergranulation vs Proliferation (HvsP) and Proliferation vs Epithelialization (PvsE). Color bar = activation z-score range. Hierarchical clustering was utilized in Ingenuity Pathway Analysis (IPA) to group pathways with similar patterns of z-score activation or inhibition across samples. **b**. Diseases and Bio Functions using IPA. Each column represents a pairwise comparison: Coagulation vs Hypergranulation (CvsH), Hypergranulation vs Proliferation (HvsP) and Proliferation vs Epithelialization (PvsE). Color bar= activation z-score. c-e. For each phase comparison, the *p*-values and the z-scores are plotted as they follow the trajectory of temporal comparisons. Each color represents a function and the different symbols represent the phase comparison: 0 CvsH, □ HvsP, 6 PvsE. **c**. Signalling pathway functions. **d**. Immune response functions. **e**. Coagulation and clotting functions. **f**. Predicted upstream regulators with columns showing their molecule type, Benjamini Hochberg corrected *p*-value for each pairwise comparison, Coagulation vs Hypergranulation (CvsH), Hypergranulation vs Proliferation (HvsP) and Proliferation vs Epithelialization (PvsE). Darker purple reflects higher significance (2 color gradient from min to max values, for white to dark purple). Activation z-score red gradient for values greater than z-score = 2 (pathway activation), blue gradient for values less than z score = −2 (pathway inhibition). Number of molecules targeted and found in the wound fluids. **g**. Skin cell types retrieved from Human Protein Atlas (HPA) analysis grouped by protein expression in the wound fluids, L=low, M=medium, H=high expression (HPA column). Significance levels are plotted as −log10(p_adj_) for all samples combined (All), Coagulation (C), Hypergranulation (H), Proliferation (P) and Epithelialization (E). Purple color gradient represents the gradient of *p*-values for All samples together, while orange color gradient represents the gradient of *p*-values for C, H, P, and E.

To identify regulator molecules that could not be detected in the wound fluids, we performed pairwise comparisons with IPA Upstream Regulators Analysis. This allowed us to predict potential biological functions that could also explain the transitions between phases (Fig. 4f). Our results show a strong enrichment of immune response and response to infection (INFG, TNF, IL6, IL1B), which show high activation scores in the transition from Coagulation to Hypergranulation, a decrease in the second transition to increase again in the transition from Proliferation to Epithelialization. A calcium channel in contractile cells, RYR1 has an increasing activation score. TGFB1 is another factor that shows with high probability to be a regulator and has been shown to be essential for regeneration^31^. Additionally, metabolism regulator insulin -like growth factor binding protein 2 (IGFBP2) is inhibited while D-Glucose appears as activated, nuclear factor erythroid 2-related factor 2 (NFE2L2), a protein that is involved in response to oxidative stress, also shows a fluctuating transition. Three growth regulators oncostatin M (OSM), MYC and tumor protein P53 (TP53), are also highly activated in the first comparison, decrease in the second and present a high activation score at the final transition. This data suggests two waves of immune response, accompanied by waves of metabolism and growth regulation, which were also observed in the diseases, function, and signaling pathways analysis (Fig. 4b-e). The targets of these molecules found in our samples and a complete list of upstream regulators are provided in Supplementary Data 9.

We then used gProfiler2 and Gene Set Enrichment Analysis (GSEA) to analyze the proteins determined for an enrichment of gene ontology terms and pathways. Only significant terms obtained from gProfiler2 were kept, and *p*-values were corrected for multiple testing according to standard methods. For the complete tables of the analysis, see Supplementary Data 10. From this analysis, we plotted the cell types associated with skin and enriched according to HPA analysis, shown in Fig. 4g. Notably, the presence of mature cells is significantly higher in the last phases, such as eccrine glands, arrector pili muscle cells, and follicle cells. Those associated with the epidermis have an increasing significance towards the last phase, such as the basal layer, corneal, granular, and spinous layers. However, those cells associated with the immune system are higher in the middle phases, such as lymphocytes, Langerhans cells. Of special attention are the fibrohistiocytic cells, a cell type usually associated with skin tumors because these cells have fibroblastic as well as macrophage-like characteristics. These results confirm that epithelialization and maturation of the skin occur at the end of regeneration. The immune and coagulation response is also prevalent across phases, and a strong antioxidative response is in place. The particular case of high activation of neutrophil degranulation could be an indicator of high antimicrobial and proteolytic proteins, which was also highlighted in the DEPs analysis. Complementary, the GSEA analysis provided a gene list with their ranked scores for each phase comparison and the gene set enriched in each phase (Supplementary Fig. 5 and Supplementary Data 11). The top-five hallmark database terms associated with Coagulation indicate a strong blood response, and ROS pathway activation, while terms in Hypergranulation indicate coagulation and an inflammatory response. The enriched terms in Proliferation and Epithelialization phases indicate a strong metabolic response. Taken together, our analyses indicate that the four phases of HFR are proteomically distinct, and they are characterized by a strong regulation of oxidative stress and a strong immune response. These features are likely the key to preventing infections while allowing for tissue remodeling and proliferation, which finally results in the restoration of tissue and function of the fingertip.

### Critical differences between HFR and animal models of regeneration

The main and critical differences between our findings and the well-studied limb regeneration (salamanders) and digit tip regeneration (mouse) are highlighted in Fig. 5. The adult Mexican salamander, the axolotl, closes the limb injury by an epithelial cell layer in the early 5 to 9 hours^32^. This epithelium matures to a specialized structure known as the apical epidermal cap (AEC), creating signaling loops that facilitate the migration and dedifferentiation of cells, resulting in a temporary structure called the blastema^33^. In parallel, histolysis remodels the skeleton while the initial cell differentiation occurs^34^. Finally, growth and differentiation progress until the limb is finally restored (differentiation and morphogenesis)^15^. In P3-segment amputated mice digit tips, regeneration follows distinct phases: starting with coagulation, immune response, and histolysis, followed by epithelial migration normally within a week, and full closure once bone histolysis allows the migration of the epithelial cells on top of the bone. Through the formation of a blastema and redifferentiation, the amputated digit tip will reform^22,24^. The early wound closure is then a common trait and extends to non-human primates, such as Rhesus monkeys, where epithelialization results in wound closure after 7 days. In our patient cohort, however, it was not possible to identify a mature wound epithelium macroscopically, and a stark difference to animal models is that the epithelialization occurs at a later phase of regeneration. While the definite presence of a blastema remains unsolved in HFR^21^, we observe high proliferation subsequent to histolysis and inflammation as suggested by the presence of mitosis and cell division markers (Supplementary Fig. 4). Despite the limitations of studying human regeneration, we show that the analysis of wound fluids is a tractable method to molecularly distinguish the processes orchestrating phases of human epimorphic regeneration.

**Fig. 5.**
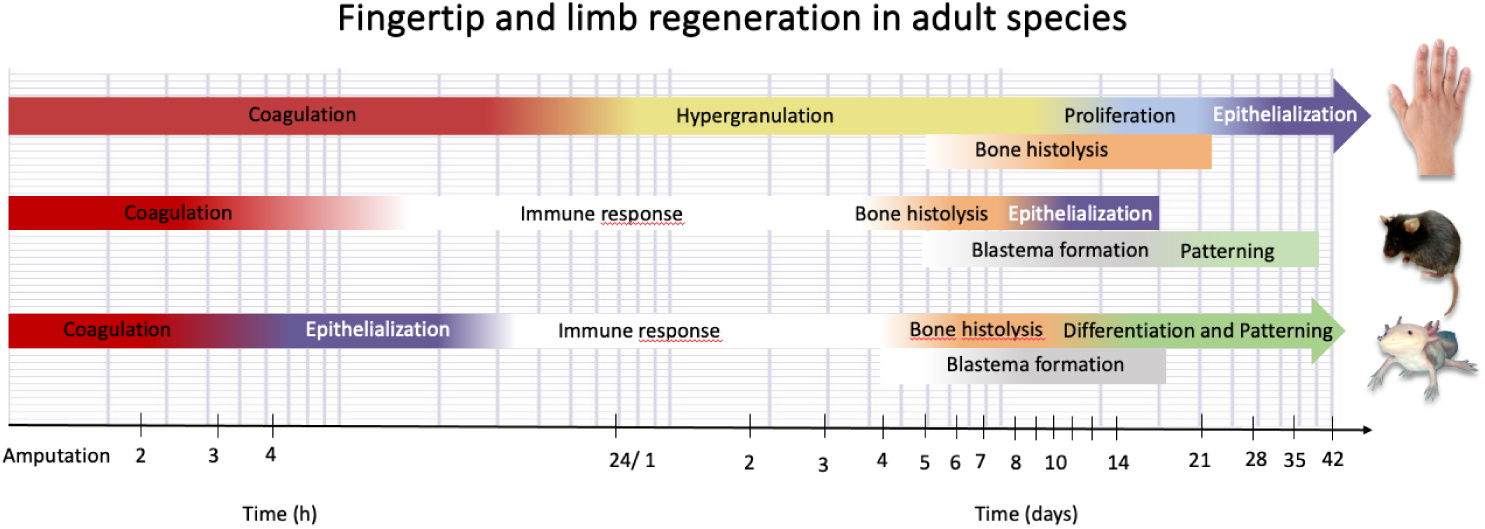
Timeline comparison of appendage regeneration in common animal models and human HFR. In salamander limb regeneration, Coagulation and Epithelization are the early steps in regeneration, occurring within 24 hours after injury in adult animals ^15,32,74,75^. In the mammalian model of digit tip regeneration, the mouse, an epithelium closes once the process of bone histolysis has been completed, followed by the formation of the blastema and cellular differentiation ^20,22,76^. In contrast, HFR is characterized by the formation of the epithelium as the last step in the process. Red in the timeline depicts the approximate duration of the Coagulation phase; yellow marks the approximate duration of Hypergranulation; Blue encompasses Proliferation and Purple Epithelialization. Similar colors label the approximate duration in animal models. Additionally, the white label indicates the inflammatory phase, the orange label represents histolysis, the gray label represents blastema formation, and the green label denotes differentiation and patterning. This representation provides a general overview and, for simplicity, does not include the overlap between phases.

## Discussion

Our work describes for the first time that human fingertip regeneration follows a sequence of distinct phases. These phases become overt in their clinical appearances, starting with the formation of a blood coagulum followed by the outgrowth of hypergranulation tissue, and a late formation of a keratinized epithelium that covers the hypergranulation tissue. The proliferation of the epithelium begins at the plane of the amputation injury and grows towards the distal apex of the hypergranulation tissue. This new tissue will become the new fingertip. When the epithelium has covered the entire hypergranulation tissue and the morphology of the fingertip is restored, many patients experience a result close to a *restitutio ad integrum*. A comprehensive quantification of aesthetic and functional results has been published previously^30^. The duration of regeneration correlates with the severity of the injury (Allen classification). The phases described here, offer a general overview of regeneration which is of interest to clinicians and patients. At the same time, the sequence of phases and their protein markers serve as an important reference in the field of regenerative biology. Molecular and cellular differences and similarities to animal models remain poorly understood. In human fingertip amputations, there is yet no definitive demonstration of a wound epithelium or a blastema formation^28,35,36^. However, in this study, we show some commonalities with the highly studied models. For example, we observe high proliferation, active ECM remodeling, an active immune response to pathogens, and differentiation of skin cell types. In addition, we observe high anti-oxidative activity in the wound fluids, similar to what has been described in animal models. Also, the excellent clinical outcomes of the patients suggest that the lack of evidence for a blastema or an AEC does not exclude similar processes occurring in HFR. A stark difference between HFR and regenerative models is the wet wound, normally considered a non-healing wound, with high levels of inflammation contributing to wound fluid production. It is important to note that in this trial, finger injuries were not subjected to disinfection, debridement, or antibiotics. Wounds were kept in their moist environment and with their bio-colonization intact.

A striking difference to animal models is the observation that in HFR, the formation of a mature epithelium is delayed while the wound remains raw-looking, suggesting a high remodeling mechanism is taking place. When analyzing the ECM components, it was surprising to see a high abundance of proteases, which usually indicates poor healing and is considered detrimental^37^. While we cannot make a direct correlation, HFR may present characteristics of a chronic wound. For instance, a chronic wound is usually diagnosed if the wound has not healed within 4 to 12 weeks. Matrix metalloproteinases like MMP9 and MMP8 have a moderate presence in the Hypergranulation and Proliferation phases. MMPs are involved in the breakdown of extracellular matrix and signaling, and regulating tissue remodeling^38^. MMP9 is particularly associated with wound healing and regeneration, and its downregulation results in delayed healing time^39^. However, the unregulated expression of MMP9 contributes to pathologies observed in wound fluids of chronic non-healing wounds^40^. Meanwhile, MMP8 is one of the predominant collagenases in wound healing and, similar to MMP9, is found in chronic wounds^41^. Furthermore, MMP8 overexpression prevents proper tissue repair^42^. Another marker of chronic wounds present in all phases of HFR is the neutrophil elastase ELANE. In our samples, however, we find a high abundance of antiproteases like A2M and the family of SERPINs, which also participate in coagulation and protection against proteolytic destruction. SERPIN deficiencies have been described in diabetic wounds^43^, and their role in wound healing is modulating proteolytic activity and inflammation^44^. The most abundant SERPIN in the samples is the protein A1, which is not usually present in skin (www.proteinatlas.org). SERPINB is found in the last two phases of HFR, a protein shown to be expressed in keratinocytes^45^. SERPIN A, C, D, E, F, and G are found in the first three phases and are almost absent in epithelialization. Other antiproteases significantly detected were trypsin inhibitors and PZP. Furthermore, coagulation proteins are abundant across phases. Together, this suggests that intense remodeling of the ECM is ongoing, even at later stages of HFR, mimicking some chronic wound features. However, an important contrast to chronic wounds is detected in the HFR wound fluids, where mitogenic and cell division control markers are prevalent in all phases with moderate abundance and a peak at the Proliferation phase. Further studies are needed to explore how a fibrotic scar is prevented in HFR and how the prominent ECM remodeling described here affects HFR.

The Coagulation phase is enriched with hemoglobins, antioxidant proteins, spectrins, and very distinctly, Ankyrin1 and SLC4A1, both proteins involved in cell motility and activation of erythrocytes. Antioxidant proteins such as PRXD 2 regulate the intracellular concentration of reactive oxygen species. Their abundance is higher in Coagulation and, to some extent, in Hypergranulation. It has been shown that a pro-regenerative environment is characterized by the resistance of cells to oxidative stress and senescence, as seen in the spiny mouse^46^. Gene sets (GS) arising from our bioinformatic analysis validate our findings regarding reactive oxygen species pathway, coagulation, peroxisomes, and heme metabolism. The Hypergranulation phase is the phase with fewer unique proteins, as its abundant proteins are either shared with the Coagulation phase or with the Proliferation phase. However, some proteins are abundant or associated with abundant proteins of this phase. These include integrin alpha-M (ITGAM), SERPINA1, SERPINA3, HPX, HP, A2M, and PZP. The GS terms associated with this phase comprise coagulation, complement, xenobiotic metabolism, and peroxisomes. The terms associated with the Proliferation phase include oxidative phosphorylation, apical junction, glycolysis, PI3K-AKT-MTOR signaling, and mitotic spindles. Representative proteins include H4 clustered histone 1 (H4C1), SERPINB10, a serine protease inhibitor. Cytokinesis proteins such as ACTB, ACTN1 and MYH9 are higher in this phase. Cell division and mitosis markers are found across the four phases but predominantly in the Proliferation phase (Supplementary Fig. 4). The Epithelialization phase is characterized by proteins associated with the keratinization processes, epithelial differentiation, and cell-cell adhesion, in particular, KERATIN type I cytoskeletal KRT10 and KRT6B. Filaggrins are proteins that aggregate keratin filaments during terminal epidermal differentiation and therefore highly specific for squamous epithelial cells and keratinocytes. Filaggrin deficiency is linked to atopic dermatitis since these proteins maintain the integrity of the skin barrier^47^. Another protein specific for this phase is desmoplakin (DSP), an important anchor protein for intercellular junctions.

One set of proteins of interest in the HFR process includes the calcium-binding proteins S100A8/A9, which are usually absent in healthy tissues^48^. These proteins have been previously associated with wound healing^49^ and promoting growth in human keratinocytes^50^. Both proteins are damage-associated molecular pattern molecules (DAMPs) and are upregulated in hyperproliferative epidermis^51^. The most abundant proteins of the family in our samples were S100A8/A9, with a peak in Proliferation. The abundance of Lactotransferrin is also of particular interest: it is abundant in all phases, with the highest abundance in the last three phases. This protein, normally secreted by macrophages, has been identified to induce a pro-regenerative phenotype in fibroblasts in the ear pinnae tissue of the spiny mouse, remaining absent in the non-regenerative ear injuries of lab mice^52^. This protein is usually not highly expressed in homeostasis (www.proteinatalas.org). The most abundant proteins in all samples are hemoglobins; the oxygen that hemoglobin carries has been deemed crucial for wound healing^53^. Here, hemoglobins show a distinct decreasing curve, although still detected in the Epithelialization phase. Similarly, the dominant plasma protein, albumin, is important for maintaining osmotic pressure in wounds^54^. In animal models, albumin administration promoted the transcription of factors related to wound healing^55^.

Additionally, our analysis of pathways and functions revealed that HFR presents strong immune and coagulation responses, which overreach the four phases. This is characterized by a strong activation of granulocytes, consistent presence of neutrophiles, as well as the innate and adaptive immune response (Fig. 2f cluster II, Supplementary Fig. 2 and Fig. 4). The transition of these functions is not linear, but rather as waves, according to the pairwise comparisons. A strong response to other organisms is clearly significant, especially because the wounds are populated by microbes, yet, infections are endogenously controlled. This is prominent in Hypergranulation and Proliferation (as seen in cluster III of Fig. 2f and Supplementary Fig. 2). A strong response to oxidative stress is also prevalent across phases. Reactive oxygen species (ROS) need to be tightly controlled during wound healing; an excess causes oxidative damage, and their absence hinders healing^56^. What remains uncertain is how the balance of ROS observed in HFR contributes to the regeneration of the fingertip, rather than wound healing, and if it plays a role in the management of infections. Furthermore, our analysis confirmed that during Epithelialization, the maturation of the epithelium and its ECM occurs. HFR involved regulated cell migration and changes in tissue texture and underlying mechanics during regeneration, indicating mechanosignaling likely contributes to regeneration. Proteome analysis of wound fluids supported this observation at the molecular level, with 14 of 60 DEP matching prior analysis of genes responding to mechanical stress, strain, pressure, or stiffness^57^. This includes molecules mentioned in mechanosignaling, electrosignaling, and mechanics of diseases contexts: ALB, CAT, F2, MPO, and PLG, as well as DEPs detected during HFR that have protein family members implicated in mechanosignaling: KRT1, SLC4A1, APOA1, APOB, SERPINA1, SERPINA3, SERPINB1, and SERPINC1. HFR DEPs highlighted mechanosignaling networks related to both free radical scavenging via CAT/SOD1 signaling and coagulation regulation anchored by PLG, APOB, and SERPIN family interactions. These new data complement observations made four decades prior, noting that mechanosensory neuron regeneration contributes to recovery of hand function after injury^58,59^.

The fingertip injuries we investigated in our RCT showed various morphological appearances^30^. Oblique cuts were difficult to classify with the existing Allen classification, especially injuries that resulted in crushed soft tissue. However, the unbiased proteomic clustering of our samples by phase makes a valid categorization according to defined clinical phases. Our MS/MS approach is not meant to provide quantitative data. We used TSC to estimate the abundance of marker proteins in our samples. Since we are the first to perform comprehensive MS/MS analysis of HFR wound fluids, this explains why we cannot compare our results to those of previous research. An internal longitudinal comparison had to compensate for the lack of standards, such as wound fluids from non-regenerating human fingertips. This internal control also had to compensate for the lack of housekeeping proteins. Lastly, one can only speculate about the role of protein degradation in wound fluids, bacterial proteins and the reliable detection of low and high-abundant proteins. In addition, fluid collection, as opposed to tissue collection, masks the role of transcription factors regulating cell dedifferentiation, reprogramming, and other mechanisms uncovered in regenerative animal models.

In conclusion, our results demonstrate that HFR has distinct phases: Coagulation, Hypergranulation, Proliferation, and Epithelialization. It is the first proteomic approach to the fascinating process of HFR and its differences from other mammalian regeneration models. We present the first comprehensive and longitudinal proteomic analysis of HFR phases from one day after injury to the end of Epithelialization. For the first time, we show that wound fluid proteomes give insights into HFR. Despite the heterogeneous cohort, a robust and unbiased separation of the regenerative phases is observed. The distinct proteomic patterns based on the phases of HFR could be demonstrated. Quantitative techniques to define key players, as well as the role of physical and electrical cues, are needed to dive deeper into the cascades that orchestrate HFR. These could then be tested in cell models. Finally, the role of bacterial colonization should be addressed in further experiments. With the experiences of human regeneration research, an important step in understanding wound healing in general will be accomplished. It remains unknown how a wound with delayed epithelialization and wound closure that presents features of a chronic wound will finally heal with minimal to no fibrosis. This study reveals the initial look into HFR at the molecular level and expands the possibilities for future therapies.

## Methods

### Sub-study to a randomized clinical trial with a novel finger cap and phase attribution

An RCT testing a novel finger cap for treating fingertip amputation injuries was conducted from September 2017 to October 2019^30^. During this trial, a novel silicone finger cap was used to occlude fingertip amputation injuries in 22 patients, 2 to 72 years old. The new device was tested in a pseudo-cross-over design. Patients were randomly assigned to start with either the silicone finger cap or a conventional film dressing. After two weeks, patients were changed to the other treatment modality. After another two weeks, patients were asked to choose between the finger cap and the film dressing for the remaining treatment^60^. Patients were not subjected to wound disinfection, wound debridement, or antibiotics. Since the novel finger cap includes a reservoir for excess wound fluid, samples of the wound fluid were aspirated with a syringe at regular intervals. Samples were stored at −80°C directly after aspiration.

To define the clinical stages of HFR, a comprehensive analysis was conducted using all available clinical notes collected during the patient visits and patient-reported outcome measures (PROM). Additionally, the regenerative progress was documented with images of the fingertip. The data was complemented with other sources, such as a previously published case series^61^ and case reports^62^. Additionally, X-rays were taken at the first visit and after a few weeks.

Fingertip amputation injuries were classified according to the Allen score. Type I involves damage only to the pulp of the finger. Type II injuries include pulp and nail loss. Type III injuries include a partial loss of the bony terminal phalanx. Type IV injuries involve the lunule, and thus, the germinative nail matrix is likely injured^63^. This grading of the amputations was done by experienced pediatric surgeons upon the first presentation after the injury and retrospectively confirmed by evaluating the radiographs and the initial photographic documentation.

Patients were included within the first 24 hours after the amputation (Visit 1). Further visits occurred the next day and weekly until the wounds were fully epithelialized (Visits 2 to 7). While patients were treated with the novel finger cap, if wound fluid was present, this was aspirated from the silicone cap and immediately frozen^60^. We used data from all 22 patients, when available, to clinically describe the regenerative phases. For the MS/MS analysis, 5 out of 22 patients were excluded for the following reasons: initially unreported diabetes mellitus (n=1), initially unreported shortening of protruding bone by primarily treating physician (n=1), superficial Allen Type II injury epithelialized within two weeks resulting in samples comprised of different phases (n=1), and insufficient wound fluid for aspiration (n=2).

### In-Gel Digestion

Gel bands (regions) were excised and cut into small cubes of approximately 1 mm side length. Gel pieces were destained and proteins were reduced, alkylated, digested in-gel with trypsin, and extracted according to standard procedures with slight modifications (digestion in 20mM NH_4_HCO_3_)^64,65^. Digests were dried in a vacuum concentrator and stored at −20°C until analysis. The complete list of materials is found in Table I.

**Table I.** SDS-PAGE Gel Electrophoresis materials.

| Item (Chemicals) | Manufacturer (Supplier) | Order Number |
| --- | --- | --- |
| BOLT™ 10% Tris-Bis Precast Gels | Invitrogen/ThermoFisherScientific |  |
| NuPAGE™ MOPS-SDS-Buffer (20x) | Invitrogen/ThermoFisherScientific | NP0001 |
| Pierce™ Lane Marker Non-Reducing Sample Buffer | ThermoFisherScientific | 39001 |
| PageRuler™ Plus Prestained Protein Ladder (10-250kDa) | ThermoScientific/ThermoFisherScientific | 26619 / 26620 |
| β-Mercaptoethanol | Merck |  |
| Mini Gel Tank | Invitrogen/ThermoFisherScientific | A25977 |

### Mass spectrometry

For LC-MS/MS analysis, dried digests were recovered with 3 µl 30% formic acid containing 50 fmol/µl isotopically labelled peptide standards (Peptide Retention Time Standards, Table II), immediately diluted with 20 µl water, and transferred to HPLC vials. For analysis, 5 µl of the sample was injected into the HPLC system. The peptide mixture was loaded onto a trap column and desalted for 10 min with solvent A (0.1% formic acid) at a flow rate of 3 µl/min. After desalting, the trap column was switched in-line with the separation column, operated at a flow rate of 200nl/min. After equilibration with solvent A, peptides were separated in a linear gradient of 90 min from 0% - 60% solvent B (60% acetonitrile, 0.1% formic acid). The mass spectrometer (Q-Exactive HF, ThermoScientific, Bremen, Germany) was operated in data-dependent (DDA) acquisition mode. Parameters of the acquisition are provided in Table III. Peptide and protein identification was performed with Mascot V3.1.0 (Matrixscience)^66^, using the database of human reviewed proteins (UP5640 reviewed only) downloaded on 14-02-2025.

**Table II.**
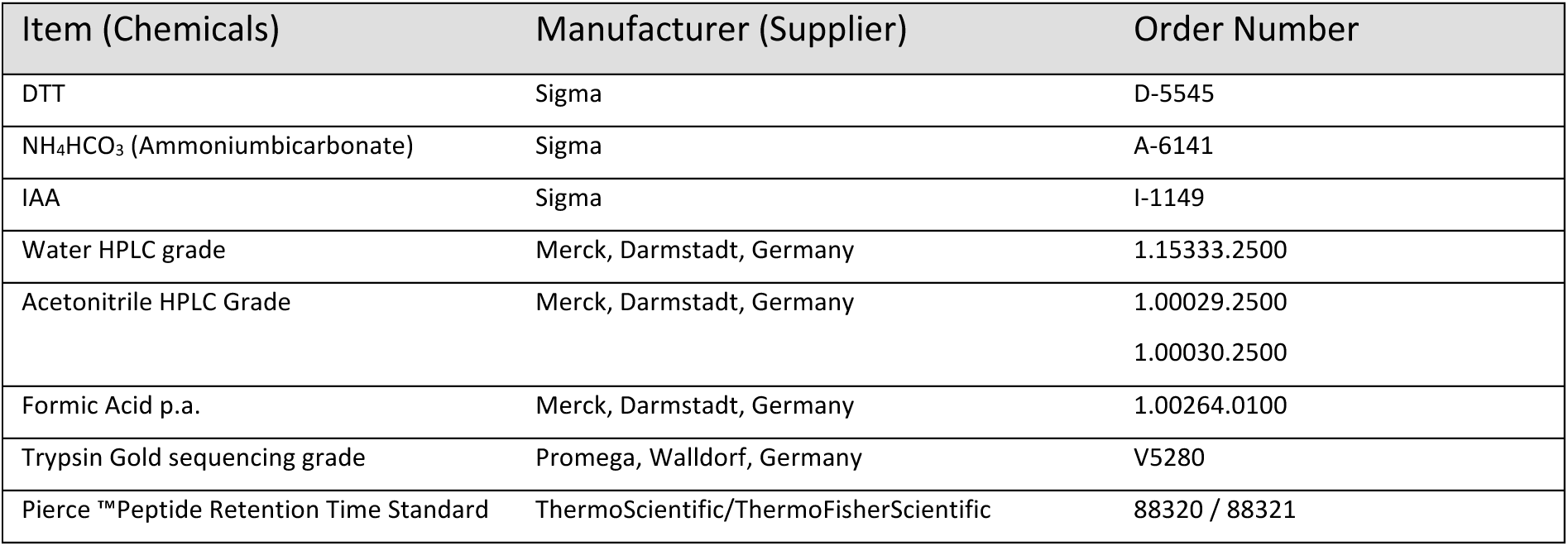
List of chemicals.

| Item (Chemicals) | Manufacturer (Supplier) | Order Number |
| --- | --- | --- |
| DTT | Sigma | D-5545 |
| NH<sub>4</sub>HCO<sub>3</sub> (Ammoniumbicarbonate) | Sigma | A-6141 |
| IAA | Sigma | I-1149 |
| Water HPLC grade | Merck, Darmstadt, Germany | 1.15333.2500 |
| Acetonitrile HPLC Grade | Merck, Darmstadt, Germany | 1.00029.2500<br>1.00030.2500 |
| Formic Acid p.a. | Merck, Darmstadt, Germany | 1.00264.0100 |
| Trypsin Gold sequencing grade | Promega, Walldorf, Germany | V5280 |
| Pierce™ Peptide Retention Time Standard | ThermoScientific/ThermoFisherScientific | 88320 / 88321 |

**Table III.**
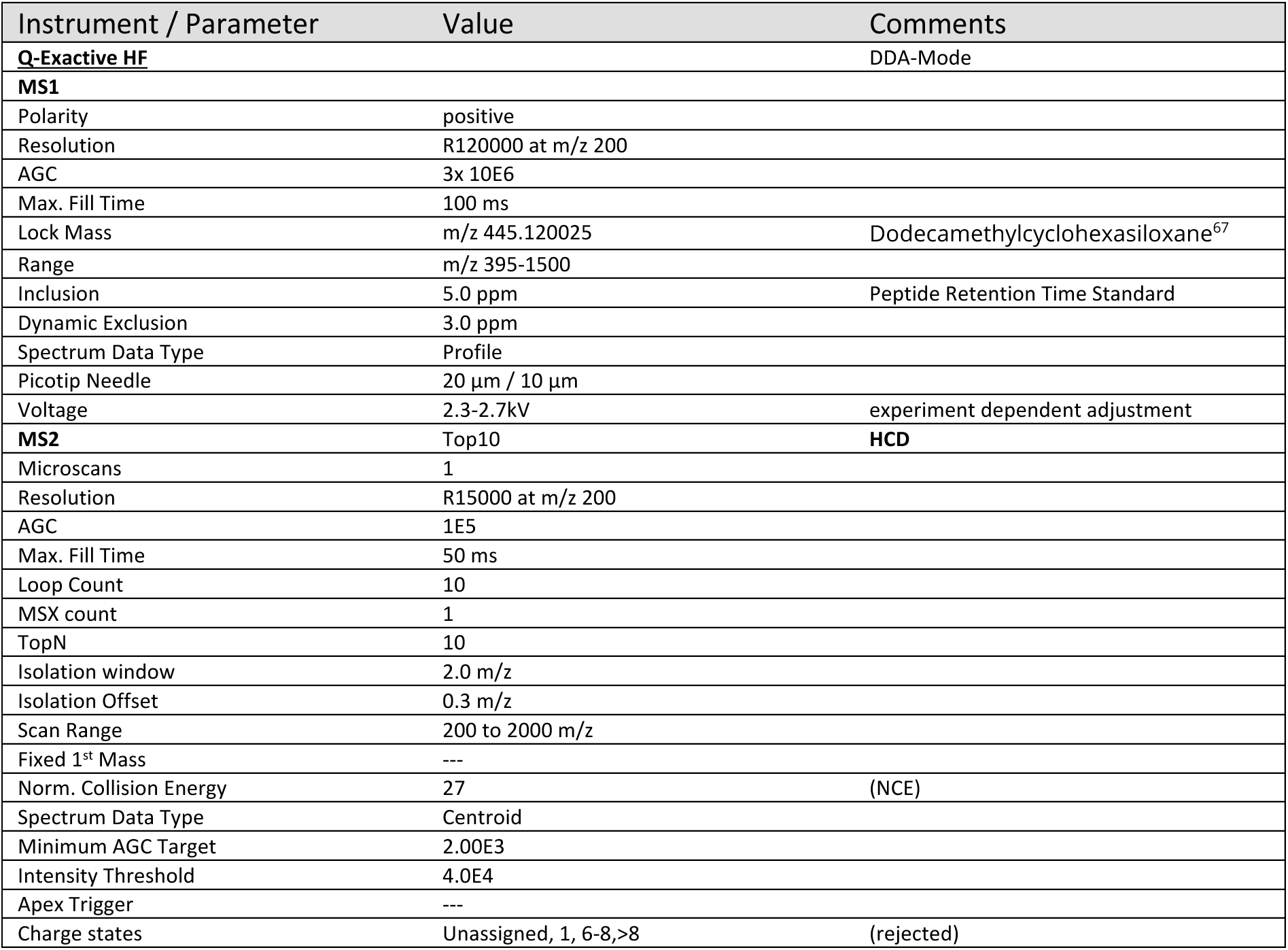
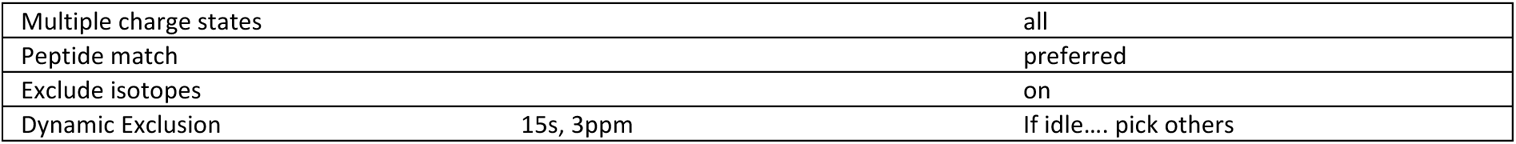
Mass Spectrometer Q-Exactive HF -DDA-.

Trypsin was set as the protease, and a maximum of 3 missed cleavages was allowed. Carbamidomethyl-cysteine was set as fixed, and methionine oxidation and protein N-terminal acetylation were set as variable modifications. Peptide hits were evaluated and aggregated in Scaffold 5.2.2 (ProteomeSoftware, USA). A detailed description of chemicals, instrumentation, and software parameters are provided in Table III, IV and V.

**Table IV.** Instrumentation specifications.

| Instrument / Material | Manufacturer (Supplier) | Comments/Order Number |
| --- | --- | --- |
| **Dionex3000 Ultimate UPLC RLSC System** | ThermoScientific, Idstein, Germany | Nanoflow System |
| Acclaim PepMap 100 C18 3µm, 2cm x 75µm,<br>Acclaim PepMap C18 2µm, 75µm x 15cm | ThermoScientific, Idstein, Germany | Trap-Column Setup<br>#164535<br>#164534 |
| Picotip Needle 10µm id | NewObjectives, Woburn, USA |  |
| **Eksigent 425 UPLC System** | Sciex, Darmstadt, Germany | Nanoflow UPLC System |
| Acclaim PepMap 100 C18, 2 µm, 75 µm x 2 cm,<br>Acclaim PepMap C18, 2 µm, 75 µm x 15 cm | ThermoScientific, Idstein, Germany | Vented-Column Setup<br>#164535<br>#164534 |
| Picotip Needle 10 µm | NewObjectives, Woburn, USA |  |

**Table V.** Software.

| Instrument / Material | Manufacturer (Supplier) | Comments |
| --- | --- | --- |
| MSConvert V3.0 <sup>68,69</sup> | Proteowizard, CDN | File Conversion Tool |
| MASCOT V3.1 | MatrixScience, London, UK | Protein Identification Software<br>matrixscience.com |
| Scaffold V5 <sup>70</sup> | Proteome Software, Portland, OR, USA | Protein Identification Statistics and Validation, Visualization<br>Proteomesoftware.com |
| Perseus 2.0.10.0<sup>71</sup> | MPI of Biochemistry, Munich, Germany | https://maxquant.net/perseus/ |

### Proteomic data analysis and statistics

#### Data Preprocessing

Total spectrum values measured for the proteins in each sample were used to create a basic common protein count data matrix. This matrix contains the corresponding unique total spectrum counts for 2481 proteins for each of the 36 wound fluid samples measured (Coagulation: 7 samples, Hypergranulation: 8 samples, Proliferation: 12 samples, Epithelialization: 9 samples). Each of the 2481 proteins was detected in at least one of the 36 patient- and healing stage-specific samples. For samples in which a specific protein was not detected, the corresponding count value was set to zero. Total spectrum values (TSV) were filtered for high confidence hits prior to numerical and proteomic analysis, specifically those mapping to the human reference proteome (Uniprot ID UP000005640) with at least 99% probability to a single protein or protein cluster (Scaffold software) and including two or more peptides mapping with at least 95% confidence. For increased stringency to analyze only true positive proteins, the 2481 proteins detected in at least 1 sample were filtered to those proteins present in any 3 or more samples, analogous to the guideline that for quantitative proteomics, proteins are detected in at least half of the measured samples. 974 proteins or protein clusters passing this filter were used for all subsequent analyses. This last filter removed ∼40% of lower-confidence entries. TSV data was analyzed using custom Python codes, PRISM 10 software (GraphPad) statistical software, and the bioinformatics tools GSEA, gProfiler and Ingenuity Pathways Analysis (IPA, Qiagen). Generative AI (ChatGPT-4) was used to speed coding for routine syntax operations.

#### Principal Component Analysis

To determine how samples relate to each other overall, principal component analysis was applied using Python library *sklearn.decomposition.PCA* using default parameter values, including centering, no scaling, and the single value decomposition solver *SciPy.linalg.svd*. PCA scores (*fit_transform; top 3 principal components shown)* and the percent explained variance for each principal component (*explained_variance_ratio * 100%; plotted as positive coefficients in green and negative in red)* were graphed for each proteome sample for the top three principal components. Average TSV per protein was also compared as a function of wound healing phase, with the most abundant proteins correlating with high magnitude coefficients in PCA analysis.

#### Clustering

Proteins were clustered for all samples, annotated by their wound healing phase, using the Seaborn library *clustermap.* Data were normalized to the maximum value per protein across all 36 samples. Zero values were replaced with 1’s to avoid division by 0. Proteins (rows) were clustered and the resulting dendrogram was used to identify main groups of proteins with similar patterns of expression across wound healing phases. Clustergrams were generated for all proteins (n=2481 proteins) using Seaborn, and using GraphPad for the smaller subset of proteins detected as significantly differently expressed (n=60 DEPs) as a function of wound regeneration phase. For the high-dimensional heatmap, the largest clusters matching distinct temporal patterns were further analyzed, showing the average spectral count per wound healing phase for all proteins within a cluster (Clusters I through V). When clustering DEPs, a small subset of the data, both protein-normalized values (Fig. 2) and raw values (Supplementary Data 4) were shown.

#### Statistical Analysis

Proteins were defined as significantly differentially expressed analyzing TSV data using two factor ANOVA (GraphPad PRISM; protein and wound phase), with Tukey’s multiple comparison post-hoc test to identify pairwise differences between wound healing phases. TSV expression values were plotted for each sample as a function of wound healing phase for all proteins (selected individual proteins included in figures).

#### Bioinformatics Analysis: Pathway and Network Analysis

To identify biological processes differentially regulated during regeneration, we manually grouped the 60 differentially expressed proteins into thematic groups based on involvement in similar biological processes (including coagulation, inflammation, extracellular matrix, and oxidative stress). Molecules from each thematic group were annotated for direct interactions within the group using IPA software (Qiagen). Network diagrams show known connections, including self-interactions and molecule types, encoded by node shape (cytokine: square, enzyme: diamond, transcriptional regulator: oval, and other: circle). Resulting network diagrams are shown for inflammation and immune signaling (12/12 DEPs directly interact) and extracellular matrix and cytoskeletal motility signaling (17/19 DEPs directly interact). Network layouts show the node with the highest connectivity in the center.

#### Term Enrichment Analysis for Biological Features

Term enrichment was determined by analyzing Entrez Gene IDs for target gene lists using gProfiler (https://biit.cs.ut.ee/gprofiler/gost) against all 3 GO ontologies (molecular function, biological process, and cellular component), KEGG and Reactome signaling databases, TRANSFAC and miRTarBase regulatory motif databases, the Human Protein Atlas and CORUM protein databases, and the HP human phenotype database. Analysis was limited to human, annotated genes. Results are shown in Fig. 4 and Supplementary Data 10. Routine Gene Set Enrichment Analysis was performed to independently confirm the predicted biological processes involved^72,73^.

#### Biological Network Regulation and Statistical Analysis

We employed the bioinformatics knowledge base and statistical analysis software Ingenuity Pathways Analysis (Qiagen) for high-throughput annotation of empirically-documented relationships between proteins expressed in wound healing phases. We obtained direct connections between the set of statistically significant, differentially expressed proteins using analysis of custom lists and permitting only direct connection annotations. For robustness of biological process interpretation, three independent Core Analyses were performed by seeding molecule lists with one of 3 sets of protein expression data: (1) significantly differentially expressed proteins (n=60 proteins), (2) all detected proteins (n=974 proteins), and (3) pairwise comparisons of expressed proteins at HFR phase (CvH, HvP, and PvE). Fold change values were used when performing core analyses and comparative analyses, to enable quantitative prediction of pathway up- or down-regulation, as well as improve statistical confidence of predicted upstream regulators. Results from comparison (1) above are shown in Supplementary Data 6. Results from comparison (2) corroborated both phase-by-phase IPA analysis, gProfiler, and GSEA analyses described above. For streamlined results visualization, comparative analysis results from analysis (3) above are shown in Fig. 4. These data include top candidates from IPA’s canonical pathway and diseases, and biological processes results. Secondary analysis of IPA results for canonical pathways enabled us to observe changes in signaling pathways activated throughout HFR. These canonical pathways were grouped into overarching functions manually and displayed as a function of their likelihood of pathway activation (or inhibition; z-score on y-axis) and the likelihood the pathway changes signaling pattern between and pair of sequential phases (shapes on plots, Fig. 4C-E; x-axis, Benjamini-Hochberg corrected *p*-values). Finally, IPA regulatory and upstream analyses tools were used to identify molecules that may regulate the observed changes in protein expression levels at each HFR phase. Tabular display of this data shows Benjamini-Hochberg corrected *p*-values to assess the significance of the upstream regulator, a z-score reflecting the magnitude and direction of fold change, and the number of target molecules (from within the maximal set of 974 proteins) detected during this phase transition and regulated by the upstream regulator. Pseudo-coloring tabular data by z-score (high to low: red to blue) shows a set of upstream regulators most upregulated in the first HFR phase transition, the intermediate phases, and a final set of regulators predicted to mediate the final phase transition between Proliferation and Epithelialization. A limited set of regulators is predicted to be downregulated across all phases. Data files showing IPA core analysis results are available as Supplementary Data 9.

## Data Availability

All data produced in the present study are available upon reasonable request to the authors

https://www.researchgate.net/publication/320317238_Study_protocol_for_a_randomized_controlled_pilot-trial_on_the_semiocclusive_treatment_of_fingertip_amputation_injuries_using_a_novel_finger_cap

## Acknowledgements

We would like to thank all members of the Sandoval-Guzmán and Doyle Labs and the Department of Pediatric Surgery, University Hospital Dresden. Grace goes to Michael Haase, who has advised on the sample storage and preparation. The RCT and the sub-study to acquire wound fluids were funded by the German Federal Government, ZIM, Grant KF3277901CR3. Parts of the processing of the MS/MS samples were funded by the Medical Faculty C. G. Carus, Technical University Dresden, MeDDrive-Grant 60481. PP and AMD were supported by the Deutsche Forschungsgemeinschaft (DFG, German Research Foundation) under Germanýs Excellence Strategy – EXC-2068 – 390729961-Cluster of Excellence Physics of Life of TU Dresden, with additional funding to AMD by the Federal Ministry of Education and Research (BMBF) and Freestate of Saxony under the Excellence Strategy of the Federal Government and the Länder. The MS/MS-analyses were funded by the DFG grant SA 3349/3-1. RA was supported by an Alexander von Humboldt-Stiftung research fellowship (PRT 1208176 HFST-P) and a Deutsche Forschungsgemeinschaft (DFG) EigeneStelle Grant (AI 214/1-1).

---

**Supplementary Fig.1.**
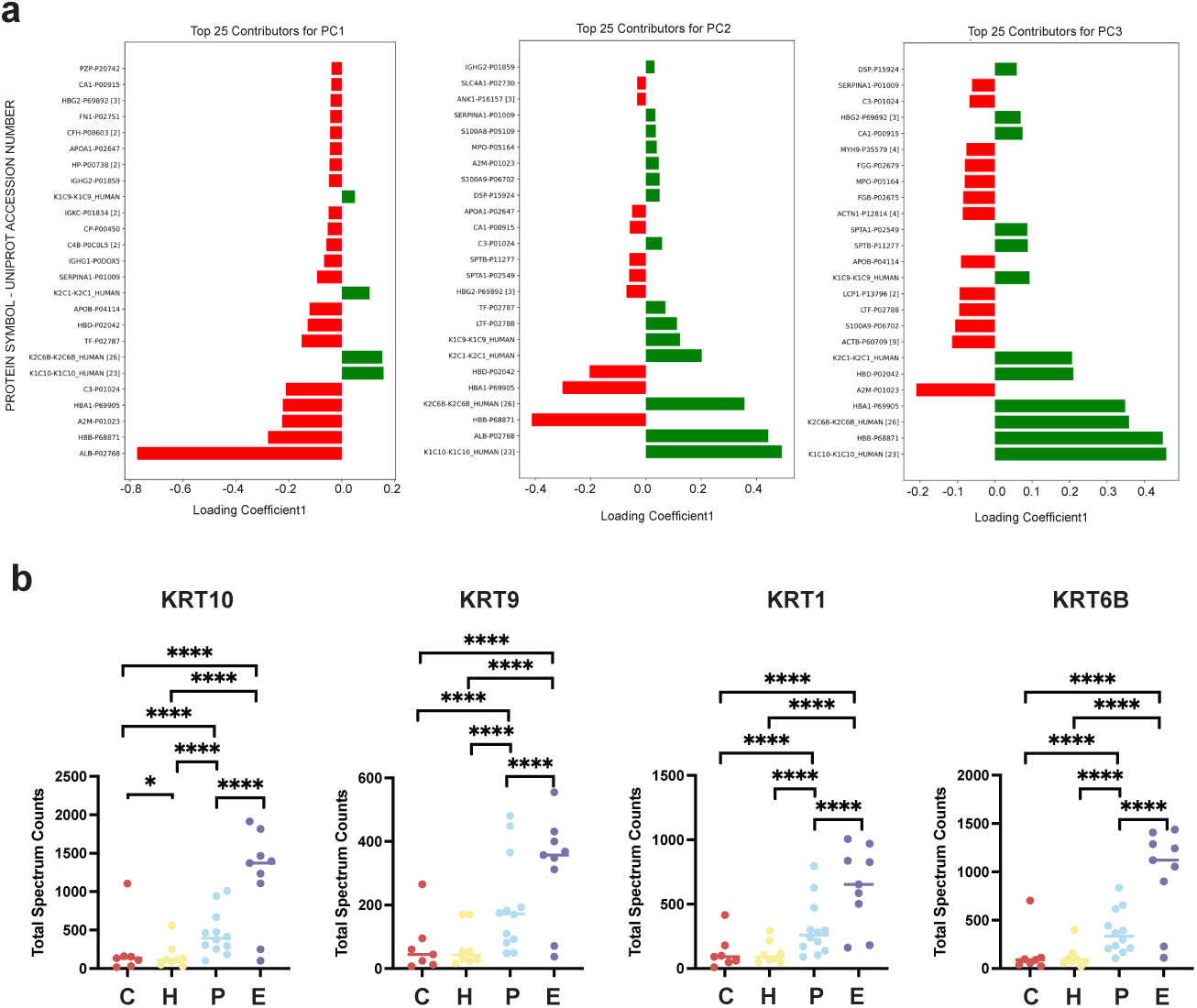
Protein contribution to PCA analysis. **a.** Top 25 protein contributors to the separation of phases in the PCA1, PCA2, and PCA3, plotted by their ID name and number, and their Loading Coefficient. Red indicates a positive coefficient contribution to the principal component, while green indicates a negative coefficient contribution to the principal component. **b.** Keratins are a primary contributor to the variation in PC3. KRT10, KRT9, KRT1 and KRT6B were detected by MS/MS analysis. One way ANOVA; significance denoted by ns (*p*>0.05), *(*p*≤0.05), **(*p*≤0.01), ***(*p*≤0.001), or ****(*p*≤0.0001).

**Supplementary Fig. 2.**
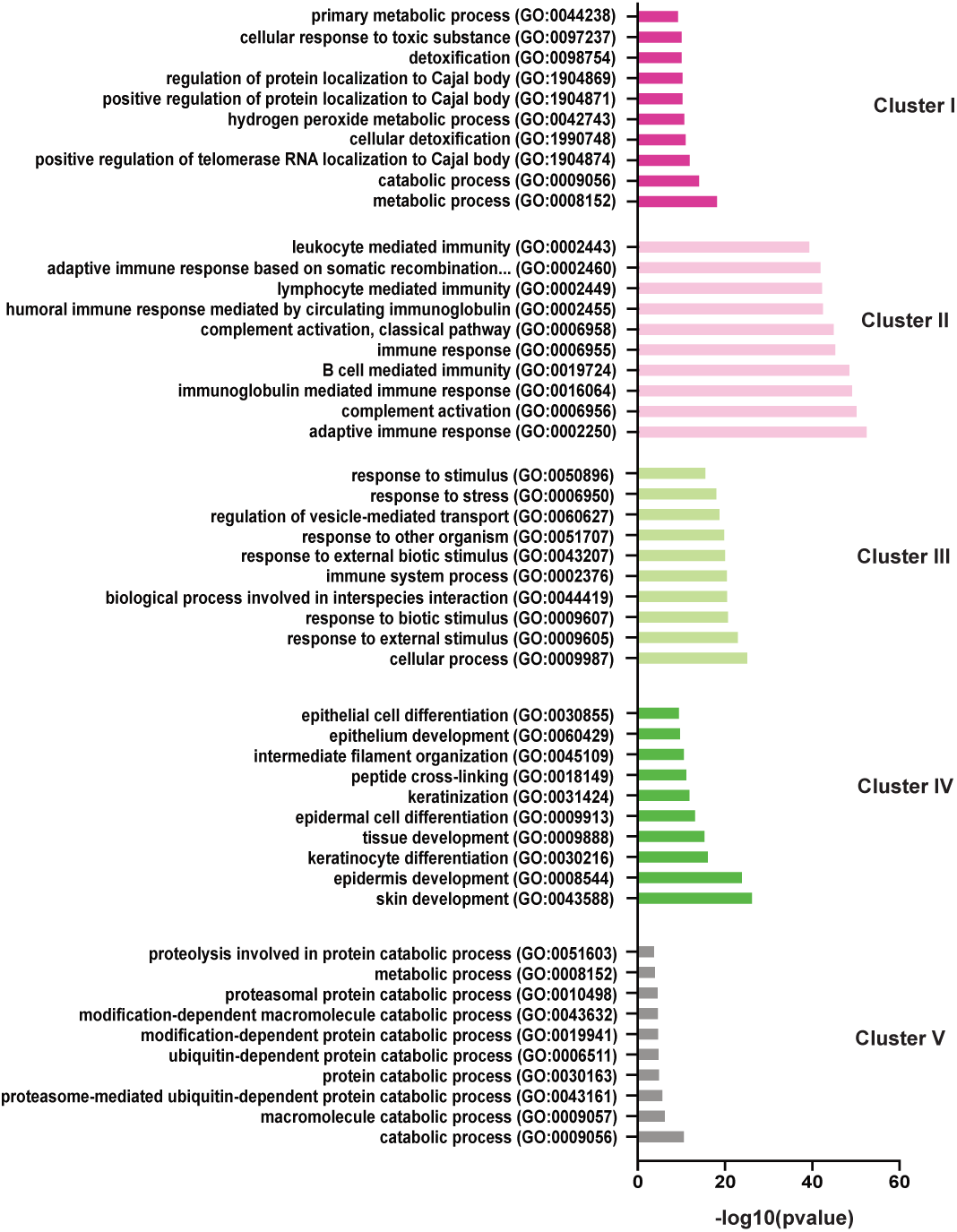
Gene Ontology (GO) enrichment analysis. Gene ontology (GO) enrichment of Biological Processes was performed using Panther, representing the Top 10 significantly enriched terms for the proteins included in the 5 sub-clusters in Fig. 2f. Terms from the category of Biological Processes were selected using Fisher’s Exact test and a Bonferroni correction.

**Supplementary Fig. 3.**
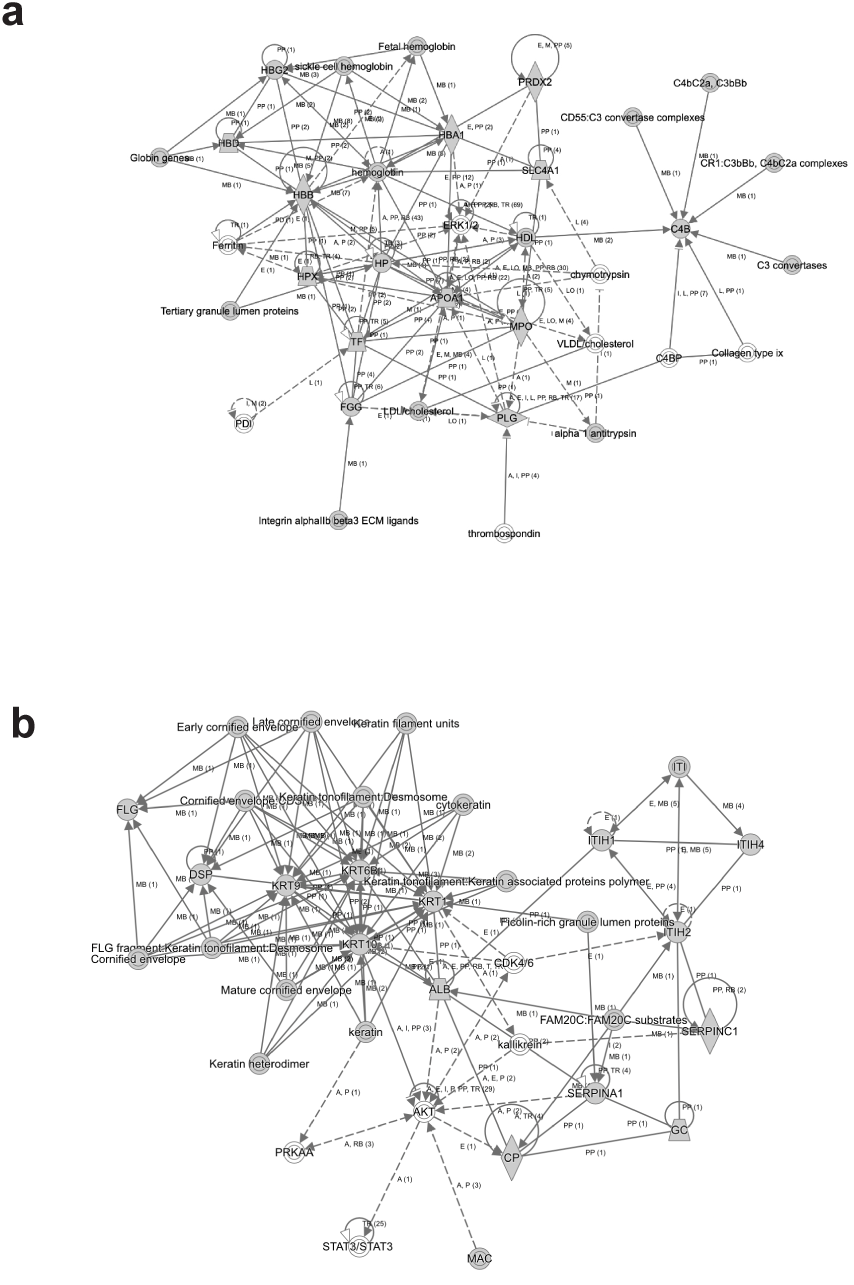
Network Diagrams of the DEPs. IPA Network diagrams showing the highest degree of direct interactions between the 60 DEPs proteins for all phases. **a.** The first network has a prominent presence of blood/immune proteins. **b.** The second network is prominent with components of ECM and its regulators. Direct and indirect interactions are represented as solid and dotted lines. Shapes represent proteins labeled by their name and their interactions supported by at least one reference from the literature (number in parentheses).

**Supplementary Fig. 4.**
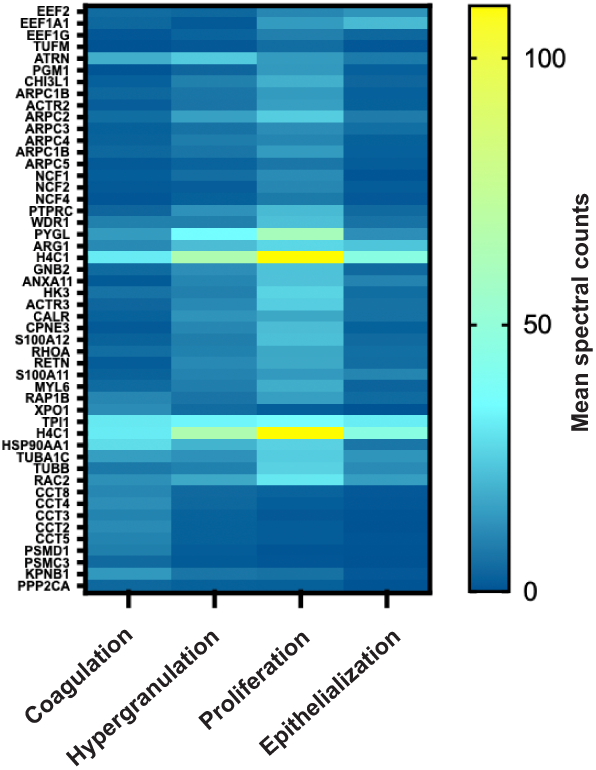
Heatmap of selected cell division and mitosis markers. Proteins are plotted as average total spectrum counts per phase. Coagulation, Hypergranulation, Proliferation, and Epithelialization.

**Supplementary Fig. 5.**
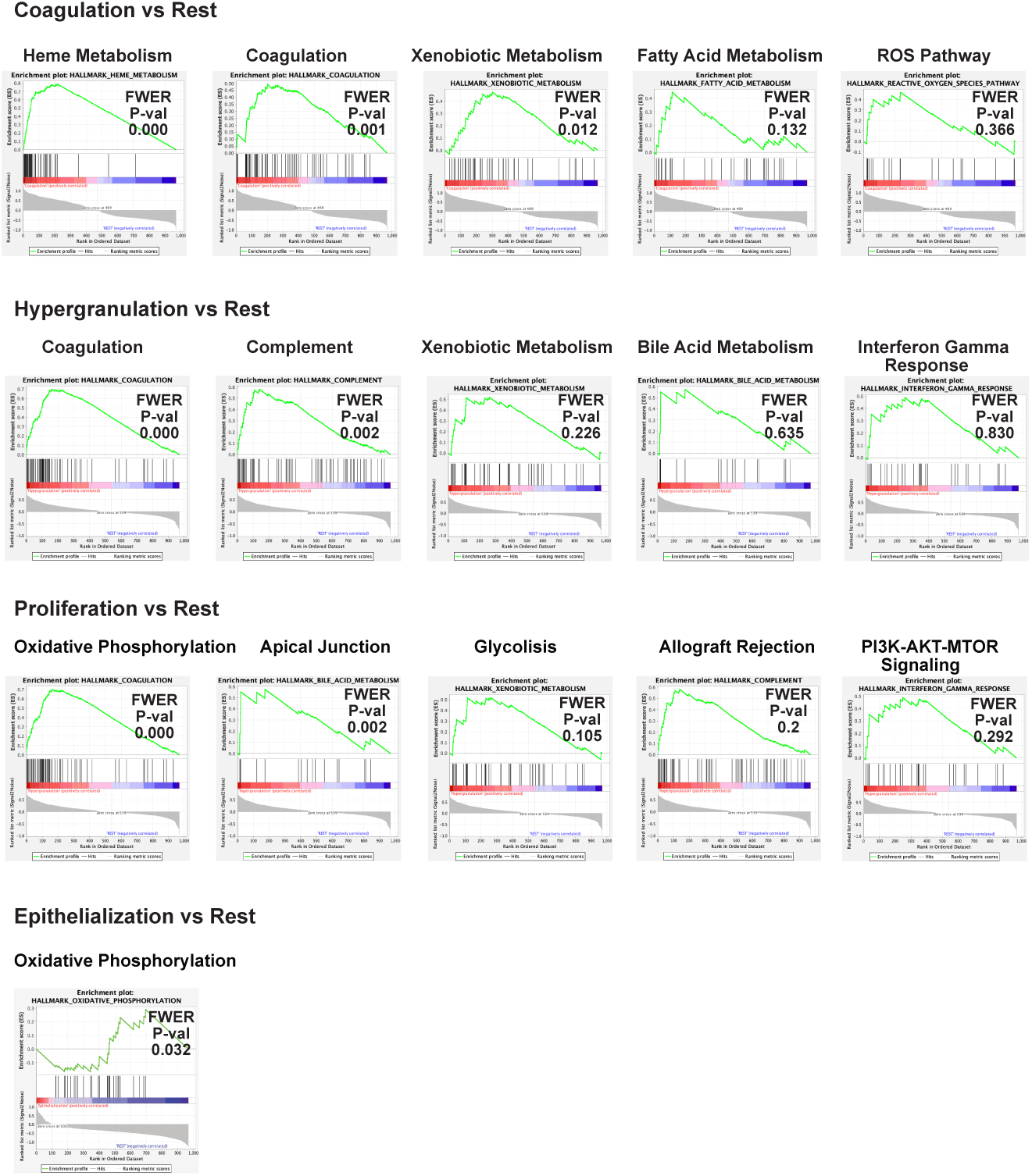
GSEA analysis with top-five hallmark terms for each phase. Coagulation, Hypergranulation, Proliferation, and Epithelialization in comparison to the other regenerative phases. Graphs depict the positive correlation in red and the negative correlation in blue. FWER *p*-value (P-val) is included for each term graph.

